# Establishment of National DRL for CT in Hybrid Imaging Studies (The Second Phase of the National NM CT (PET) Dose Audit for Kuwait Population -2019)

**DOI:** 10.1101/2020.09.20.20198176

**Authors:** Michael Masoomi, Iman Al-Shammeri, Latifah Al-Kandari, Hany Elrahman, Jehan Al-Shammari

**Author notes:** **Corresponding Author** Dr Michael Masoomi, FIPEM(UK), MRCP(Lon), SRCS(UK), Nuclear Medicine &Molecular Imaging Unit, Department of Medical Imaging, Adan Hospital, MOH, 46020, Kuwait. Participating Centers: 1. Adan Hospital – KW 2. Farwaniya Hospital - KW 3. Al-Jaber Hospital - KW 4. Chest Diseases Hospital –KW 5. Mubarak Hospital -KW 6. Al-Jahra Hospital – KW 7. Kuwait Cancer Control Center – KW 8. Jaber Al Ahmed Molecular Imaging/KFAS – KW.

## Abstract

DRLs for CT part used in PET/CT examinations are limited. The aim was to execute the second phase of the national DRL for CT part of PET/CT imaging operating in KW, in support of optimisation and dose reduction as imaging technology is advancing. In this multicentre collaborative PET centers (No:8) audit, data collection was restricted to the adult oncology patients due to a limited number of the other studies and also due to the National MOH Ethical Committee recommendation. The CTDIvol, DLP and SL were recorded and the Median, Mean, SD, 75^th^, 25^th^ percentiles as well as WB effective dose (ED) were calculated. Dose and scan length statistics for HB and WB scans (65% and 35% of total: 309) and the WB+HB presented together with the proposed NDRLs and the Achievable doses. Third quartile DLP (mGy x cm) and CTDIvol (mGy) values for the HB were (537, 5) which were higher than the UK NDRL (400, 4.3) but were lower than the Swiss NDRL (620, 6) and the France NDRL (762, 7.7). Comparatively, the Proposed NDRLs for (WB) were (684, 4.1) which were lower than Swiss National Data (720, 5.0). It is worth noted that, the Swiss had about 5000 (HB) & 706 (WB), the UK had 370 (HB) and France had 1000 (HB) entries. Calculated ED varied from 4.1 to 10.2 mSv, (mean values=6.9 mSv) for HB and from 2.6 to 7 mSv (mean value=4.6 mSv) for WB scans which were lower than the first phase (2018). Although, there was 9.1% improvement in NDRL, but the outcome suggested there is a continuous need for monitoring NDRL.

## INTRODUCTION

As we stated in the first phase of this study [**1**], there is an increased global focus on the need to carefully manage radiation exposures from CT imaging, as the doses from the related examinations are in general, higher than those from most other medical x-ray imaging examinations.

Recent development of PET/CT systems has allowed concurrent or with minimum time delay to perform anatomic and functional imaging of organs, which allows better evaluation of disease [**2-4**]. The CT components of PET/CT systems are equivalent in power output to their stand-alone versions and may be used for diagnostic purposes under appropriate CT technique. In addition to diagnostics acquisition, the application of CT in hybrid PET/CT scanners may serve other specific purposes, including attenuation correction of the PET image data and tissue localization [**5**]. The CT dose measurement concept is based on the CT dose index (CTDI), which represents the average absorbed dose of irradiation of contiguous slices.

An ^18^F-FDG PET/CT examination has potential for high patient dose from the combination of the x-rays used to acquire the CT image and the radiotracer administered into the patient to acquire the PET image. The methodology to estimate the radiation exposure of patients receiving ^18^F-FDG has been well studied and understood [**6**]. Generally, administered ^18^F-FDG activities range from 1.25 to 5 MBq/kg, depending on the sensitivity of the PET scanner (2 or 3-dimensional) [**7**], which equates to a whole-body effective dose of approximately 8.4 mSv for a 70-kg patient who receives an activity concentration of 5 MBq/kg. Including CT as part of a PET/CT examination can raise the total effective dose to as high as approximately 26 mSv [**8-11**]. However, estimates of PET/CT effective dose are variable and critically depend on an institution’s specific CT technique and activity administration protocol [**12**]. In general, the CT component of a PET/CT examination can contribute more than 50% of the total-examination effective dose.

On contrary to diagnostic CT, published dose reference levels for CT used in hybrid PET/CT examinations and guidance on CT dosimetry metrics in the literature on nuclear medicine practice standards are limited, and many of available reports reference to dedicated diagnostic CT practice standards, which may not be appropriate for CT in PET/CT. Of the limited documentation on CT technique for PET oncology, there is a general acceptance that CT dose is tailored to its purpose in the reconstruction or the interpretation process [**12-13**]. For example, acquisition protocols may be designed to compensate for the PET temporal resolution [**14-15**] or collect ultra-low-dose attenuation correction data [**16**]. There are a limited number of studies investigating the optimisation of CT technique and image quality for diagnostic and oncologic PET/CT. Most of these studies have focused on lowering estimates of effective dose while maintaining high image quality using the Alderson RANDO anthropomorphic phantom (The Phantom Laboratory) [**8-9 & 17**].

An internationally recognized approach to radiation protection of patients, recommended by the International Commission on Radiological Protection (ICRP), is the establishment and use of diagnostic reference levels (DRLs) which are dosimetric indicators, established from surveys of imaging practice and provide guidance to help manage dose and promote optimization, so that the applied dose is appropriate for a given clinical need. Furthermore, a widely accepted approach to optimization of medical radiation exposures, recommended by the ICRP [**18-19**] and the International Atomic Energy Agency (IAEA) [**20**], is the establishment and use of national, regional and local DRLs. Diagnostic reference levels (DRL) of the volumetric CT dose index (CTDI_vol_) which are levels for Whole-Body/Half – Body CT used in PET/CT examinations are limited [**2**].

The substantial variations in dose, from imaging procedures, between some health care facilities for the same examination or procedure and similar patient group (adults or children of defined sizes) have been noticed following the related. Such observations indicate the need for standardization of dose and reduction in variation in dose without compromising the clinical purpose of each examination or procedure. Examination-specific or procedure-specific DRLs for various patient groups can provide the stimulus for monitoring practice to promote improvements in patient protection [**21**].

DRLs should be set for representative examinations or procedures performed in the local area, country or region where they are applied. National DRLs (NDRLs) should be set on the basis of wide scale surveys of the median doses representing typical practice for a patient group (e.g. adults or children of different sizes) at a range of representative healthcare facilities for a specific type of examination or procedure. NDRLs are commonly set at the third quartile values (the values that splits off the highest 25% of data from the remaining 75%) of these national distributions [**22**]. As such, NDRLs are not optimum doses, but nevertheless they are helpful in identifying potentially unusual practice, where median doses are among the highest 25% of the national dose distribution. DRLs can be also established for a region within the country or, in some cases, regions of several countries. They can be also used to set updated values for new imaging technologies that may allow lower dose levels to be achieved. Where no national or regional DRLs are available, DRLs can be set based on local dosimetry or practice data, or can be based on published values that are appropriate for the local circumstances. Clinical protocols for performing a particular examination or procedure should be reviewed if the comparison shows that the facility’s typical dose exceeds the DRL, or that the facility’s typical dose is substantially below the DRL and it is evident that the exposures are not producing images of diagnostic usefulness or are not yielding the expected medical benefit to the patient. The resulting actions aimed at improving the optimization of protection and safety, which usually, but not necessarily, result in lower typical doses of the facilities for the examinations or the procedures. The examinations or procedures included should represent at least the most frequent examinations performed in the region for which dose assessment is practicable, with priority given to those that result in the highest patient radiation dose.

There is no preferred custodian: what is important is that a patient dose database (for DRLs) is established and maintained, DRL values are set, disseminated through the regulatory processes, and a process for periodic review is established. Fast moving technological developments in medical imaging are providing new opportunities to automatically track and benchmark patient doses. Early evidence in some countries with more advanced electronic systems is very promising.

In this study, DRLs for each center was calculated based on the local practice and then a national DRL (NDRL) for oncology examinations (majority of studies) and patients group were proposed adopting the third quartile value of the volume computed tomography dose index (CTDIvol) and dose length product (DLP) following the wide scale national surveys according to the IPEM-UK frame work and the ICRP recommendation for the state of Kuwait.

A comparative study to this project were a national survey conducted by the UK (47 PET/CT centers), Swiss (16 PET/CT centers) and France (56 PET/ centers) on PET/CT oncologic procedures.

## METHODOLOGY

In this continuing study (second phase) that was a multicentre collaborative research, we collected a multiple data from CT of PET/CT hybrid imaging system (that were in current practice in Kuwait Hospitals) and analysed the results for setting up a NDRL base line for the State of Kuwait. The data collection was restricted to adult patients as per the Kuwait Ethical Committee recommendation. The Methodology, based on the UK - IPEM was adopted as per the first year audit to suit the proposed study involving the KW NM Clinical Centers. The studies were carried out with participation of 8 PET/CT centers in the state of Kuwait. WE managed to collect 309 patient data for CT part of PET/CT centers in comparison to 197 patients for the first year. The focus was on all PET/CT imaging systems and procedures regardless of their locations and numbers of the systems availability in one center.

To reduce the influence from centers that provided a significantly large number of entries compared to others, the limiting data contribution from each center was set to a maximum of 40 entries. Majority of PET/CT imagings were dedicated to the oncology scan and the collected data were based mostly on this type of imaging and the other isolated studies including heart and brain were excluded. In addition, all topograms (scanograms) and monitoring steps used in contrast-enhanced CT acquisitions (if any) were excluded from the analysis. All proposed CTDI*vol* were approximated to the first decimal place and the DLP to the nearest whole number.

Initially, a body phantom study was performed on a local GE PET/CT 710 to determine the CTDI_vol_ and DLP for an anthropomorphic phantom, and to assess feasibility of recording preliminary data for the above. The priority was to estimate typical patient dose quantities for the common present practice on adult patients according to the following steps:

- To record displayed values of radiation dose quantity for maximum samples of 40 typical adult patients per center (i.e. 309 sample data for all the centers), undergoing procedures for common clinical indications. For CT part, the clinical purpose of exposure, attenuation correction (AC), localization or diagnosis for the PET/CT imaging centers were recorded for Whole body and Half Body.
- To calculate, for each type of examination, the median values of dose quantities (e.g. CTDI_vol_ and DLP for CT); which were the typical dose levels (but were not the local DRLs that are set for a group of imaging systems or a group of hospitals).
- To compare the typical dose levels (median values) with the published DRLs for a similar practice in the absence of local or national DRLs (UK, Swiss and France), in order to provide a broad indication of our relative performance and urgency of need for improvement in our imaging technique. A proposed national DRL for Kuwait was then suggested.
- The effective dose for the oncology examination at each center, using CT part of PET/CT was calculated based on the measured dose values.
- The Sante DICOM viewer was used to retrieve all the listed parameters in the data sheets (Appendix 1).

Statistical analysis was performed on the data collected for the oncology protocol from the participating centers, in respect of volumetric CT dose index (CTDI*vol*), dose length product (DLP) and scan length (SL), taking into consideration the intended aim (attenuation correction and localization) declared by each nuclear medicine center. For the each of these metrics, the number of entries, median, mean, standard deviation, minimum and maximum values and 75^th^ & 25^th^ percentiles of the combined were calculated. Rounded third quartile values of CTDIvol and DLP were used to produce suggested NDRLs, whereas rounded first quartile values were used to produce achievable doses as a further aid to optimization [**22**-**23**]. In order to provide verification, the analyses were performed independently. Once the data were analyzed, NDRLs representing the 75th% percentile of the data distribution, were proposed in addition to achievable dose (defined as the 25th% percentile of the data distribution) for each protocol. The ICRP suggests taking the third quartile of the distribution of individual median values as the DRL. However, in this study, we present both mean and median for DRL to accommodate suggestions by the various groups, including the UK, Swiss and France national surveys. All the 8 centers in KW, except one, used Automatic Exposure Control (AEC) that modulates radiation exposure automatically and is widely used for optimization of radiation dose in CT [**24-26**].

We also estimated the effective dose (ED) as a prerequisite for optimization and monitoring of radiation exposure of the CT part of PET/CT facilities. ED is often estimated as a product of the DLP value and a conversion factor selected according to the imaging region [**27-29**]. CTDIvol is calculated on the basis of radiation dose measured in imaging 16-cm and 32-cm CT dosimetry phantoms for head-mode and body-mode imaging, respectively. When using the same scanner parameters, CTDIvol, and consequently DLP, is larger for a 16-cm phantom than for a 32-cm phantom because of less absorption within a smaller phantom. The conversion factor from DLP to ED depends on the location, size, and radiosensitivity of organs and tissues exposed to radiation and is lower for the head than for the trunk. For 18F-FDG PET/CT oncology applications, CT images are usually acquired from the head to the proximal thigh sequentially, and a single DLP value, representing half body radiation exposure, is provided on a scanner.

It is also important to note that whereas the fundamental concept of ED has not changed with new ICRP recommendations, important aspects of its calculation have been updated, leading in particular to changes in values of dose per unit exposure since the previous UK CT survey for 2003. Furthermore, ICRP has now recommended the application of specific reference persons for the calculation of the organ doses necessary for the estimation of ED using Monte Carlo techniques to simulate radiation transport in computational anthropomorphic phantoms. The ICRP adult male (AM) and adult female (AF) voxel phantoms will be adopted in future ICRP publications in place of the physical or mathematical phantoms used previously for the calculation of ED.

To assess the radiation dose from the CT component of the examination, we used dose-length product (DLP) values from the scanner-generated dose reports and a conversion factor—that is, the region-specific normalized effective dose per DLP (mSv × mGy^−1^ × cm^−1^) [**30**]. Effective dose (*ED*) was then estimated as the product of the *DLP* and the corresponding conversion factor (*k*): *ED* (mSv) ≈ *k* × *DLP*.

For the half body and whole body scan, we used a **k** value of 0.015 mSv × mGy^−1^ × cm^−1^ and 0.0093 mSv × mGy^−1^ × cm^−1^ respectively [**31-32**]. The coefficients of ED/DLP for examinations were for adult patients and were calculated as mean values, over a range of CT scanner models operating at medium applied potentials (principally 120 kVp), on the basis of ICRP 103 tissue weighting factors and ICRP 110 voxel phantoms (as an average for AM and AF) [**33**].

For the calculation of displaying CTDIvol and DLP; GE scanners (GE, Milwaukee, Wisconsin, USA) using 32 cm body CTDI phantom on all the systems in this study. Patients were not categorized by age, sex, or weight because the scanning protocol was used with the automatic exposure control (AEC), which accounted for differences in patient size.

All PET/CT centers in this study, except one, used the Adaptive Statistical Iterative Reconstruction (ASiR) that is the first commercially available reconstruction algorithm that provided significant dose benefit for CT imaging. ASiR has been accepted by numerous sites as the standard-of-care protocol for a variety of applications and has potential to achieve significant reductions in patient radiation dose in CT exams while achieving image reconstruction speed similar to that of conventional analytical reconstruction using filtered back projection (FBP).

### International comparison

The “mean and median” values of dose quantities (CTDI_vol_ and DLP for CT) for data, collected from the Kuwait multiple centers to establish whether they are above or below the published DRL. The “mean” values had been recommended earlier, but the recent recommendations are favoring median values [**34-36**].

A similar work by the CT working groups in the UK, Swiss and France have recently been performed and the results have been published [**37-39**]. Some of the protocols exercised in Kuwait are common and thus DRL’s can be directly compared. For comparison purposes, CTDI*vol* was the most relevant metric, whereas DLP depends directly on the scan length applied at individual centers.

## RESULTS AND DISCUSSION

This study has been able to generate data from a truly representative cross section of Kuwait PET/CT practice that is mostly exercised for oncology examinations. The data from other studies, including heart and brain were limited, to provide a statically acceptable and accurate results and as such were not included in this study. These studies may be considered in the future audit.

The half-body (chest, abdomen and pelvis) fluorine-18-fluorodeoxyglucose (18F-FDG) oncology imaging comprised the majority of PET/CT imaging procedures performed in PET/CT centers in Kuwait (65% of total collected data for 2019 verses 53% for the earlier audit in 2018), though there was much variation in half body studies in the centers. The rest of PET/CT studies (35% in 2019, verses 47% in 2018) performed as the whole body examination (head to toe) using F18-FDG or F18-NAF.

All the participating centers, except one, used AEC and ASiR in acquiring CT part of the PET / CT examination. Minimum and maximum range of mA for setting AEC were very much variable (14-209 mA & 81-400 mA respectively) for both HB and WB oncology PET/CT examinations. One center who did not use AER, had set the mA low, in the range of 50-83 mA for PET/CT studies, that could be due to intention to use the CT scan for the purpose of attenuation correction at the foremost. There was not much variation in setting CT tube voltage (i.e. 120kV) for the PET/CT examinations across the 8 centers and in particular all centers adopted the stated value.

Maximum variation for the “mean” SL value (cm) between 8 PET/CT centers was 9% for HB and 11% for WB PET/CT examinations according to the **Tables** [**1-2**] data. The “mean” and “median” SL values (cm) for the HB scan were (106, 106) and for the WB were (168, 168) which appeared to be higher than the UK HB (95, 94) and the Swiss HB (94, 101) and the Swiss WB (119, 128). It is worth noted that the HB was defined to include chest-abdomen-pelvis scan length and the WB scan length was referred to head to toe scan for KW protocols that could be different in setting SL from UK and Swiss protocol point of views. The Swiss had only 6 entries for WB scan whereas KW had 200 entries. The average male and female lengths for UK and Swiss national were not known.

Summary of dose and scan length statistics for the half body, whole body and the combined scan length are presented in **Tables [1-2]**, whereas **Table [3]** presents the same data for the combined half and total body examinations.

In all cases, the CT data were used for AC and localization, but acquisition parameters and patient doses for the 8 PET/CT center varied, with a maximum of two and half fold variation in the dose - length product (DLP) between centers. The third quartile of CTDIvol and DLP values were used to propose the local DRL (LDRL) and the first quartile of CTDIvol and DLP values were calculated to suggest the achievable LDRL for each participating center accordingly [**Tables 4-6**]. There was a maximum of twofold variation in LDRL for CTDIvol and DLP between seven centers [Tables 4-6]. The proposed national diagnostic reference level (NDRL) and achievable DRL (based on the median and mean values of CTDIvol and DLP) for HB, WB and HB+WB for the oncology examinations were calculated and presented in **Tables [7-8]**.

**Table 1:** Summary statistics for the distribution of the scanner volume computed tomography dose index and dose-length product for the protocol list (HB) of each centre using PET/CT.

| Center | Protocol<br>[Use: AC-L] | CTDI<sub>vol</sub> [mGy] |  |  |  |  | DLP [mGy x cm] |  |  |  |  | Scan Length [cm] |  |  |  |  |
| --- | --- | --- | --- | --- | --- | --- | --- | --- | --- | --- | --- | --- | --- | --- | --- | --- |
|  |  | Median | Mean | STD | Min | Max | Median | Mean | STD | Min | Max | Median | Mean | STD | Min | Max |
| 1 | PET Oncology<br>[HB]: 25N | 4.0 | 4 | 1.0 | 1.6 | 7.9 | 448 | 432 | 141 | 125 | 908 | 109 | 107 | 17 | 74 | 135 |
| 2 | PET Oncology<br>[HB]: 25N | 4.5 | 5 | 1.0 | 3.7 | 8.4 | 513 | 530 | 101 | 387 | 969 | 109 | 107 | 5 | 98 | 121 |
| 3 | PET Oncology<br>[HB]: 29N | 5.5 | 5 | 1.9 | 1.6 | 7.8 | 516 | 560 | 225 | 163 | 905 | 98 | 103 | 6 | 97 | 121 |
| 4 | PET Oncology<br>[HB]: 18N | 6.0 | 6 | 1.6 | 2.7 | 9.0 | 678 | 699 | 217 | 301 | 1125 | 109 | 112 | 11 | 97 | 133 |
| 5 | PET Oncology<br>[HB]: 28N | 4.6 | 5 | 1.4 | 3.3 | 8.4 | 483 | 523 | 140 | 374 | 868 | 97 | 102 | 8 | 97 | 135 |
| 6 | PET Oncology<br>[HB]: 24N | 2.9 | 3 | 0.5 | 1.8 | 3.1 | 314 | 299 | 63 | 170 | 388 | 110 | 111 | 13 | 66 | 130 |
| 7 | PET Oncology<br>[HB]: 32N | 2.6 | 3 | 0.8 | 1.4 | 4.6 | 273 | 286 | 82 | 144 | 472 | 98 | 102 | 8 | 86 | 130 |
| 8 | PET Oncology<br>[HB]: 19N | 4.0 | 4 | 1.3 | 2.4 | 7.0 | 441 | 452 | 143 | 242 | 718 | 102 | 107 | 7 | 102 | 126 |

AC- Attenuation Correction, L- Localization, HB – Half Body, N – Number of entry, CTDI<sub>vol</sub> – CT Dose Volume, DLP – Dose Length Product

**Table 2:** Summary statistics for the distribution of the scanner volume computed tomography dose index and dose-length product for the protocol list (WB) of each centre using PET/CT.

| Center | Protocol<br>[Use: AC-L] | CTDI<sub>vol</sub> [mGy] |  |  |  |  | DLP [mGy x cm] |  |  |  |  | Scan Length [cm] |  |  |  |  |
| --- | --- | --- | --- | --- | --- | --- | --- | --- | --- | --- | --- | --- | --- | --- | --- | --- |
|  |  | Median | Mean | STD | Min | Max | Median | Mean | STD | Min | Max | Median | Mean | STD | Min | Max |
| 1 | PET Oncology<br>[WB] : 15N | 2.6 | 2.8 | 0.6 | 1.8 | 3.8 | 462 | 475 | 119 | 272 | 664 | 168 | 169 | 12 | 135 | 180 |
| 2 | PET Oncology<br>[WB]: 6N | 3.0 | 2.9 | 0.6 | 2.3 | 3.5 | 432 | 441 | 157 | 101 | 611 | 168 | 166 | 8 | 156 | 180 |
| 3 | PET Oncology<br>[WB]: 11N | 2.8 | 4.2 | 2.2 | 2.2 | 8.7 | 474 | 691 | 364 | 385 | 1501 | 173 | 171 | 10 | 156 | 180 |
| 4 | PET Oncology<br>[WB]: 22N | 4.5 | 4.8 | 2.0 | 1.3 | 9.0 | 758 | 817 | 342 | 227 | 1657 | 168 | 166 | 9 | 156 | 180 |
| 5 | PET Oncology<br>[WB]: 12N | 3.7 | 3.9 | 1.4 | 2.4 | 6.1 | 607 | 664 | 243 | 424 | 1108 | 168 | 171 | 7 | 156 | 180 |
| 6 | PET Oncology<br>[WB]: 14N | 2.7 | 2.6 | 0.4 | 1.8 | 3.1 | 460 | 445 | 85 | 270 | 558 | 170 | 168 | 19 | 110 | 186 |
| 7 | PET Oncology<br>[WB]: 8N | 3.0 | 3.0 | 1.1 | 1.4 | 5.0 | 277 | 329 | 133 | 144 | 666 | 160 | 158 | 23 | 109 | 185 |
| 8 | PET Oncology<br>[WB]: 21N | 3.2 | 3.6 | 1.2 | 2.0 | 7.0 | 512 | 531 | 159 | 242 | 957 | 184 | 178 | 8 | 161 | 185 |

AC- Attenuation Correction, L- Localization, HB – Half Body, N – Number of entry , CTDI<sub>vol</sub> – CT Dose Volume, DLP – Dose Length Product

**Table 3:** Summary statistics for the distribution of the scanner volume computed tomography dose index and dose-length product for the protocol list (WB+HB) of each centre using PET/CT.

| Center | Protocol<br>[Use: AC&L] | CTDI<sub>vol</sub> [mGy] |  |  |  |  | DLP [mGy x cm] |  |  |  |  | Scan Length [cm] |  |  |  |  |
| --- | --- | --- | --- | --- | --- | --- | --- | --- | --- | --- | --- | --- | --- | --- | --- | --- |
|  |  | Median | Mean | STD | Min | Max | Median | Mean | STD | Min | Max | Median | Mean | STD | Min | Max |
| 1 | PET Oncology<br>[WB+HB]: 40N | 3.6 | 3.6 | 1.1 | 1.6 | 7.9 | 453 | 448 | 103 | 125 | 908 | 125 | 130 | 34 | 74 | 180 |
| 2 | PET Oncology<br>[WB+HB]:31N | 4.4 | 4.4 | 1.2 | 2.3 | 8.4 | 513 | 510 | 122 | 101 | 969 | 109 | 118 | 24 | 98 | 180 |
| 3 | PET Oncology<br>[WB+HB]:40N | 4.7 | 5.0 | 2.1 | 1.6 | 8.7 | 506 | 596 | 277 | 163 | 1501 | 109 | 122 | 31 | 97 | 180 |
| 4 | PET Oncology<br>[WB+HB]:40N | 4.8 | 5.3 | 1.9 | 1.3 | 9.0 | 711 | 764 | 299 | 227 | 1657 | 156 | 141 | 28 | 97 | 180 |
| 5 | PET Oncology<br>[WB+HB]:40N | 4.1 | 4.6 | 1.5 | 2.4 | 8.4 | 493 | 568 | 191 | 374 | 1108 | 109 | 123 | 33 | 97 | 180 |
| 6 | PET Oncology<br>[WB+HB]:38N | 2.9 | 2.6 | 0.5 | 1.8 | 3.1 | 339 | 353 | 101 | 170 | 558 | 115 | 131 | 31 | 66 | 186 |
| 7 | PET Oncology<br>[WB+HB]:40N | 2.7 | 2.7 | 0.9 | 1.4 | 5.0 | 277 | 306 | 110 | 144 | 666 | 99 | 113 | 26 | 86 | 185 |
| 8 | PET Oncology<br>[WB+HB]:40N | 3.4 | 3.8 | 1.3 | 2.0 | 7.0 | 490 | 496 | 157 | 242 | 957 | 161 | 144 | 36 | 102 | 185 |

AC- Attenuation Correction, L- Localization, TB – Total Body, HB – Half Body, N – Number of entry, CTDI<sub>vol</sub> – CT Dose Volume, DLP – Dose Length Product

**Table 4:** Proposed and achievable LDRL for AC and Localization product for the clinical NM examination protocol (HB) at each centre using PET/CT.

| Center | Protocol<br>[Use: AC&L] | Proposed LDRL [75th%] |  | Achievable DRL [25th%] |  |
| --- | --- | --- | --- | --- | --- |
|  |  | CTDIvol [mGy] | DLP [mGy x cm] | CTDIvol [mGy] | DLP [mGy x cm] |
| 1 | PET Oncology<br>[HB] : 25N | 4.5 | 483 | 3.6 | 366 |
| 2 | PET Oncology<br>[HB]: 25N | 4.8 | 556 | 4.4 | 493 |
| 3 | PET Oncology<br>[HB]: 29N | 7.1 | 778 | 3.8 | 368 |
| 4 | PET Oncology<br>[HB]: 18N | 7.4 | 872 | 4.7 | 573 |
| 5 | PET Oncology<br>[HB]: 28N | 4.9 | 520 | 4.0 | 433 |
| 6 | PET Oncology<br>[HB]: 24N | 2.9 | 339 | 2.4 | 256 |
| 7 | PET Oncology<br>[HB]: 32N | 3.0 | 316 | 2.1 | 223 |
| 8 | PET Oncology<br>[HB]: 19N | 5.0 | 507 | 3.3 | 349 |

AC- Attenuation Correction, L– Localization, HB – Half Body, CTDIvol – CT Dose Volume, DLP – Dose Length Product, LDRL – Local Dose reference Level, DLP – Dose Reference Level

Third quartile DLP (mGy x cm) and CTDIvol (mGy) values (537, 5) related to the Kuwait HB PET/CT scans (for setting NDRL) were higher than the current UK NDRL (400, 4.3) but lower than the Swiss National NDRL (620, 6) and the France National NDRL (762, 7.7). Comparatively, the Proposed NDRLs for (WB) was (684, 4.1) which was lower than Swiss National Data (720, 5.0). The Kuwait results were in reasonable agreement for the second consecutive year with the centers having 200 (HB) and 109 (WB) entries, where Swiss had about 5000 (HB) & 706 (WB), the UK had 370 (HB) and France had 1000 (HB) entries [**Tables 9-10**]. The calculated ED varied from 4.1 to 10.2 mSv, with a mean value equal to 6.9 mSv, for HB and from 2.6 to 7 mSv, with a mean value equal to 4.6 mSv, for WB scans [**Table 11**].

**Table 5:** Proposed and achievable LDRL for AC and Localization product for the clinical NM examination protocol (WB) at each centre using PET/CT.

| Center | Protocol<br>[Use: AC&L] | Proposed LDRL [75th%] |  | Achievable DRL [25th%] |  |
| --- | --- | --- | --- | --- | --- |
|  |  | CTDIvol [mGy] | DLP [mGy x cm] | CTDIvol [mGy] | DLP [mGy x cm] |
| 1 | PET Oncology<br>[WB] : 15N | 3.2 | 572 | 2.4 | 398 |
| 2 | PET Oncology<br>[WB]: 6N | 3.5 | 556 | 2.4 | 417 |
| 3 | PET Oncology<br>[WB]: 11N | 5.5 | 896 | 2.5 | 421 |
| 4 | PET Oncology<br>[WB]: 22N | 5.1 | 866 | 3.7 | 651 |
| 5 | PET Oncology<br>[WB]: 12N | 5.0 | 693 | 2.5 | 463 |
| 6 | PET Oncology<br>[WB]: 14N | 2.9 | 516 | 2.4 | 406 |
| 7 | PET Oncology<br>[WB]: 8N | 3.4 | 418 | 2.2 | 229 |
| 8 | PET Oncology<br>[WB]: 21N | 4.3 | 585 | 2.8 | 423 |

AC- Attenuation Correction, L- Localization, HB – Half Body, CTDIvol – CT Dose Volume, DLP – Dose Length Product, LDRL – Local Dose reference Level, DLP – Dose Reference Level

**Table 6:** Proposed and achievable LDRL for AC and Localization product for the clinical NM examination protocol (WB+HB) at each centre using PET/CT.

| Center | Protocol<br>[Use: AC&L] | Proposed LDRL [75th%] |  | Achievable DRL [25th%] |  |
| --- | --- | --- | --- | --- | --- |
|  |  | CTDIvol [mGy] | DLP [mGy xCm] | CTDIvol [mGy] | DLP [mGy xCm] |
| 1 | PET Oncology<br>[WB+HB]: 40N | 4.2 | 520 | 2.9 | 382 |
| 2 | PET Oncology<br>[WB+HB]:31N | 4.8 | 556 | 4.1 | 471 |
| 3 | PET Oncology<br>[WB+HB]:40N | 7.0 | 792 | 2.9 | 406 |
| 4 | PET Oncology<br>[WB+HB]:40N | 6.9 | 886 | 4.2 | 601 |
| 5 | PET Oncology<br>[WB+HB]:40N | 5.0 | 607 | 3.8 | 448 |
| 6 | PET Oncology<br>[WB+HB]:38N | 2.9 | 417 | 2.4 | 280 |
| 7 | PET Oncology<br>[WB+HB]:40N | 3.1 | 366 | 2.1 | 223 |
| 8 | PET Oncology<br>[WB+HB]:40N | 4.4 | 571 | 3.0 | 389 |

AC- Attenuation Correction, L- Localization, HB – Half Body, TB – Total Body, CTDIvol – CT Dose Volume, DLP – Dose Length Product LDRL – Local Dose reference Level, DLP – Dose Reference Level

**Table 7:** Proposed and achievable NDRL for AC and Localization product for the suggested clinical NM protocols using PET/CT: (Based on Mean Value).

| Center | Protocol<br>[Use: AC&L] | Proposed NDRL [75th%] |  | Achievable DRL [25th%] |  |
| --- | --- | --- | --- | --- | --- |
|  |  | CTDIvol [mGy] | DLP [mGy x Cm] | CTDIvol [mGy] | DLP [mGy x Cm] |
| **1** | PET Oncology<br>[HB]:200N | 5 | 537 | 3.8 | 398 |
| **2** | PET Oncology<br>[WB]:109N | 4.1 | 684 | 2.9 | 444 |
| **3** | PET Oncology<br>[WB +HB]:309N | 4.7 | 575 | 3.4 | 424 |

AC- Attenuation Correction, L- Localization, HB – Half Body, CTDIvol – CT Dose Volume, DLP – Dose Length Product, NDRL – National Diagnostic Reference Level, DLP – Dose Reference Level

**Table 8:** Proposed and achievable NDRL for AC and Localization product for the suggested clinical NM protocols using PET/CT: (Based on Median Value).

| Center | Protocol<br>[Use: AC&L] | Proposed NDRL [75th%] |  | Achievable DRL [25th%] |  |
| --- | --- | --- | --- | --- | --- |
|  |  | CTDvol [mGy] | DLP [mGy x Cm] | CTDvol [mGy] | DLP [mGy x Cm] |
| **1** | PET Oncology<br>[HB]:200N | 4.8 | 514 | 3.7 | 409 |
| **2** | PET Oncology<br>[WB]:109N | 3.6 | 536 | 2.7 | 444 |
| **3** | PET Oncology<br>[WB +HB]:309N | 4.5 | 507 | 3.3 | 424 |

AC- Attenuation Correction, L– Localization, HB – Half Body, CTDIvol – CT Dose Volume, DLP – Dose Length Product, NDRL – National Diagnostic Reference Level, DLP – Dose Reference Level

**Table 9:** Comparison of Proposed NDRL for AC and Localization product for the suggested clinical NM protocols using PET/CT: (Based on Mean Values).

| Center | Protocol<br>[Use: AC&L] | UK<br>Proposed NDRL |  | SWISS<br>Proposed NDRL |  | FRANCE<br>Proposed NDRL |  | KUWAIT<br>Proposed NDRL |  |
| --- | --- | --- | --- | --- | --- | --- | --- | --- | --- |
|  |  | CTDIvol<br>[mGy] | DLP<br>[mGy x Cm] | CTDIvol<br>[mGy] | DLP<br>[mGy x Cm] | CTDIvol<br>[mGy] | DLP<br>[mGy x Cm] | CTDIvol<br>[mGy] | DLP<br>[mGy x Cm] |
| 1 | PET Oncology<br>[HB]:200N | 4.3 | 400 | 6.0 | 620 | 6.6 | 628 | 5 | 537 |
| 2 | PET Oncology<br>[WB]:109N |  |  | 5.0 | 720 | 7.7 | 762 | 4.1 | 684 |
| 3 | PET Oncology<br>[WB +HB]:309N | X | X | X | X | X | X | 4.7 | 575 |

AC- Attenuation Correction, L- Localization, HB – Half Body, CTDvol – CT Dose Volume, DLP – Dose Length Product, NDRL – National Diagnostic Reference Level, DLP – Dose Reference Level

**Table 10:** Comparison of Proposed NDRL for AC and Localization product for the suggested clinical NM protocol using PET/CT: (Based on Median Values: KW only).

| Center | Protocol<br>[Use: AC&L] | UK<br>Proposed NDRL |  | SWISS<br>Proposed NDRL |  | FRANCE<br>Proposed NDRL |  | KKUWAIT<br>Proposed NDRL |  |
| --- | --- | --- | --- | --- | --- | --- | --- | --- | --- |
|  |  | CTDIvol<br>[mGy] | DLP<br>[mGy x Cm] | CTDIvol<br>[mGy] | DLP<br>[mGy x Cm] | CTDIvol<br>[mGy] | DLP<br>[mGy x Cm] | CTDIvol<br>[mGy] | DLP<br>[mGy x |
| Cm] |  |  |  |  |  |  |  |  |  |
| 1 | PET Oncology<br>[HB]:200N | 4.3 | 400 | 6.0 | 620 | 6.6 | 628 | 4.8 | 514 |
| 2 | PET Oncology<br>[WB]:109N |  |  | 5.0 | 720 | 7.7 | 762 | 3.6 | 536 |
| 3 | PET Oncology<br>[WB +HB]:309N | X | X | x | x | X | X | 4.5 | 507 |

AC- Attenuation Correction, L– Localization, HB – Half Body, CTDIvol – CT Dose Volume, DLP – Dose Length Product, NDRL – National Diagnostic Reference Level, DLP – Dose Reference Level

**Table 11:**
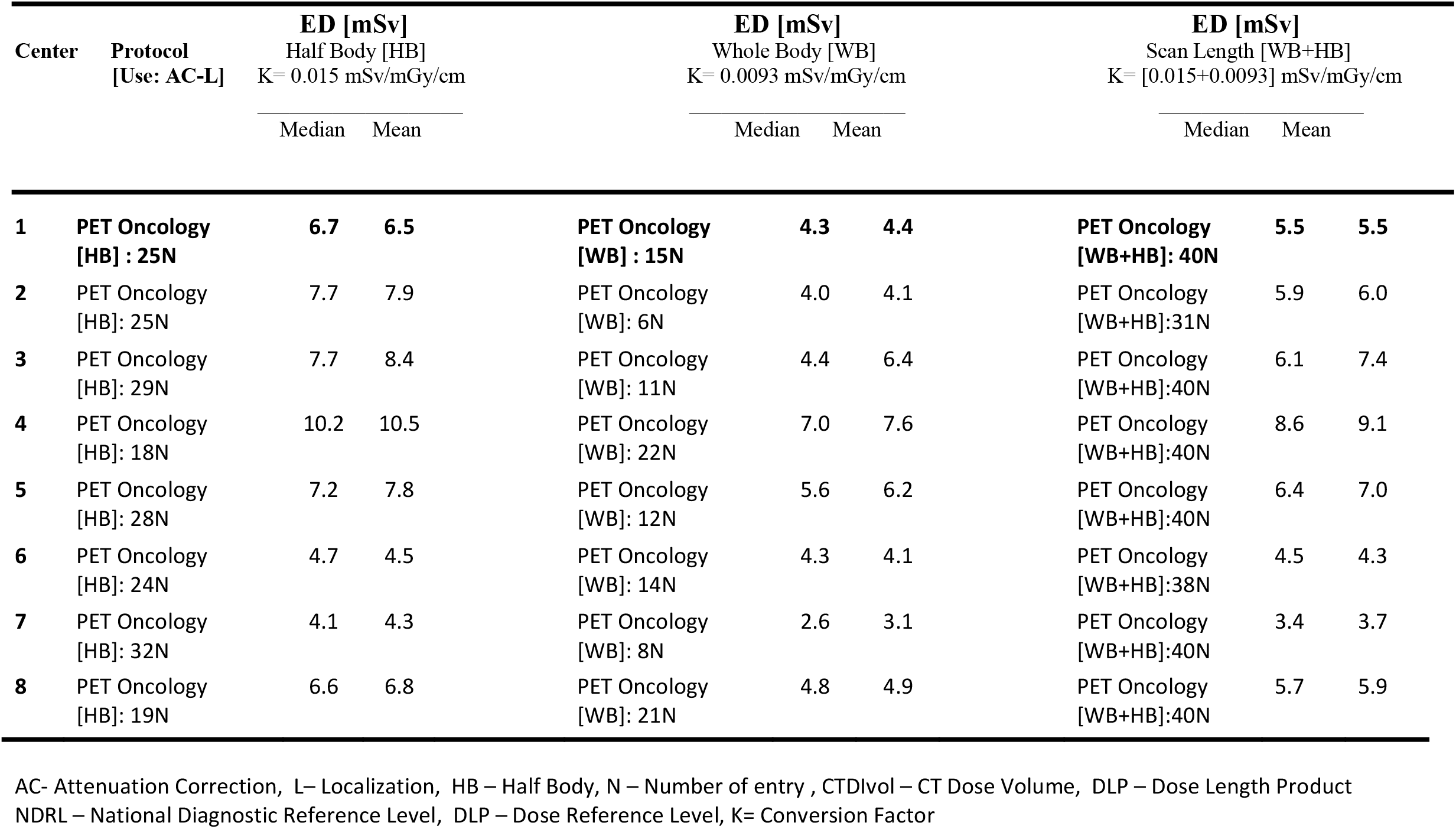
Comparison of CT effective dose as a result of AC and Localization product for the suggested clinical NM protocol using PET/CT: The recommended published conversion factors; K=0.015 (mSv/mGy/cm) & K=0.0093 (mSv/mGy/cm).

Seven out of eight centers had accommodated Discovery™ GE PET/CT with Optima™, 64 slices CT part, including two digital GE PET/CT. The remaining scanner was a Fillips Gemini PET/CT. Seven out of eight sites were reported using automatic current modulation (AEC) which goal is to automate the adjustment of tube current (mA) along the way, based on patient thickness and the radiation attenuation of tissues, patient length and the asymmetry along the patient’s body.

Data presented in **Figures [1,3 & 5]** show the range of doses (75^th^ percentile CTDIvol) for the proposed HB, WB and HB+WB oncology examinations related to AC & Localization clinical purposes. Similarly, **Figures [2,4 & 6]** are presenting variations of DLP (75^th^ percentile) for each NM center for HB, WB and WB+HB in relation to the proposed DLP for each NM PET/CT center. The dose results (CTDIvol) for 6 centers appeared to be less for the WB oncology examinations. The dose results (CTDIvol) for 6 centers appeared to be less than the proposed NDRL for HB and the WB oncology examinations. It is worth noted that two of the centers had accommodated a state of art digital PET/CT which has elevated technology other than the rest of PET/CT respectively and one center had accommodated a Philips PET/CT of an older model, setting a low mA for purpose of attenuation correction primarily, and with no use of AEC or ASiR.

**Figure 1:**
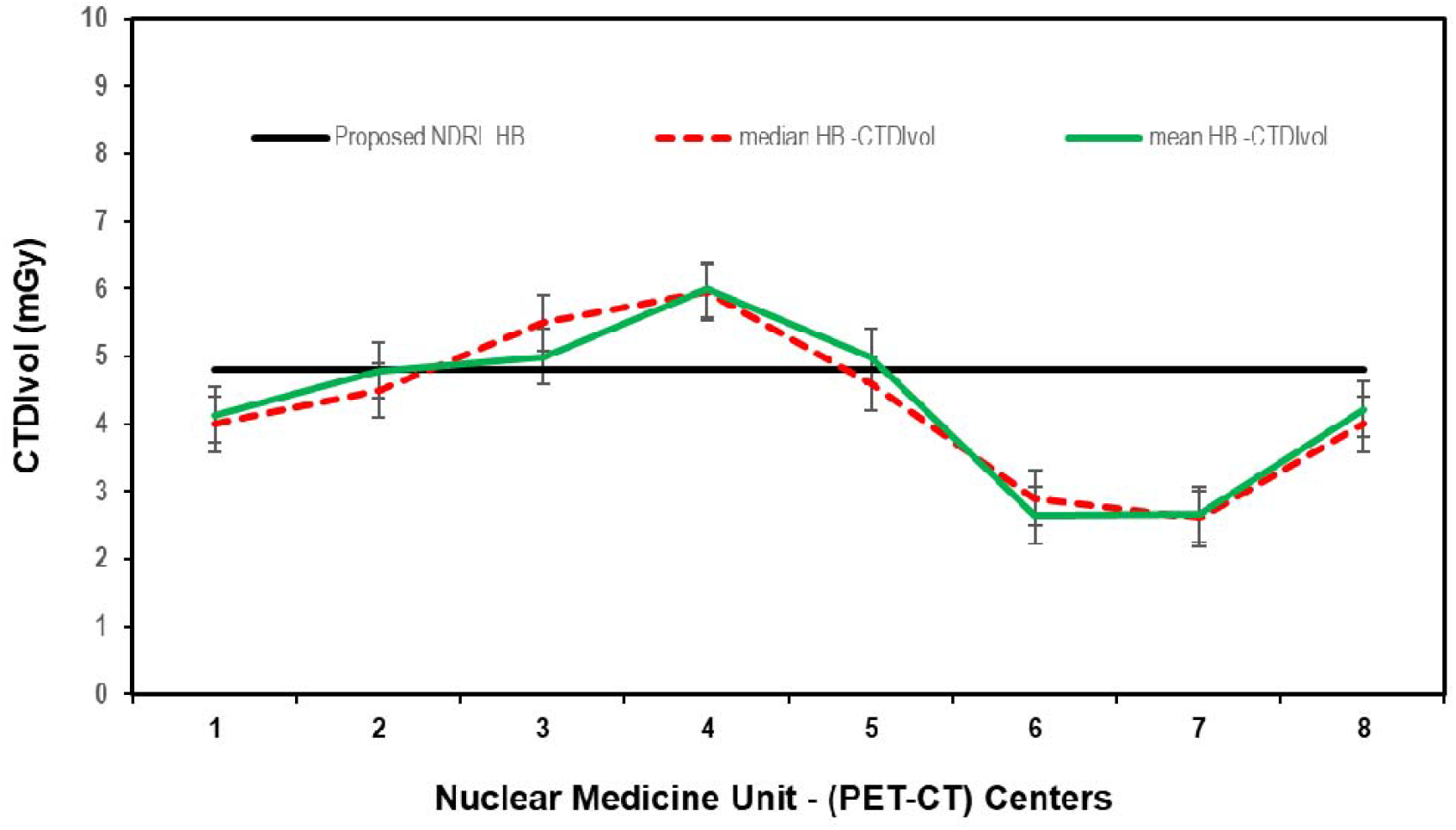
CTDIvol (75^th^ percentile) for each PET-CT unit, compared to the proposed NDRL for HB data.

**Figure 2.**
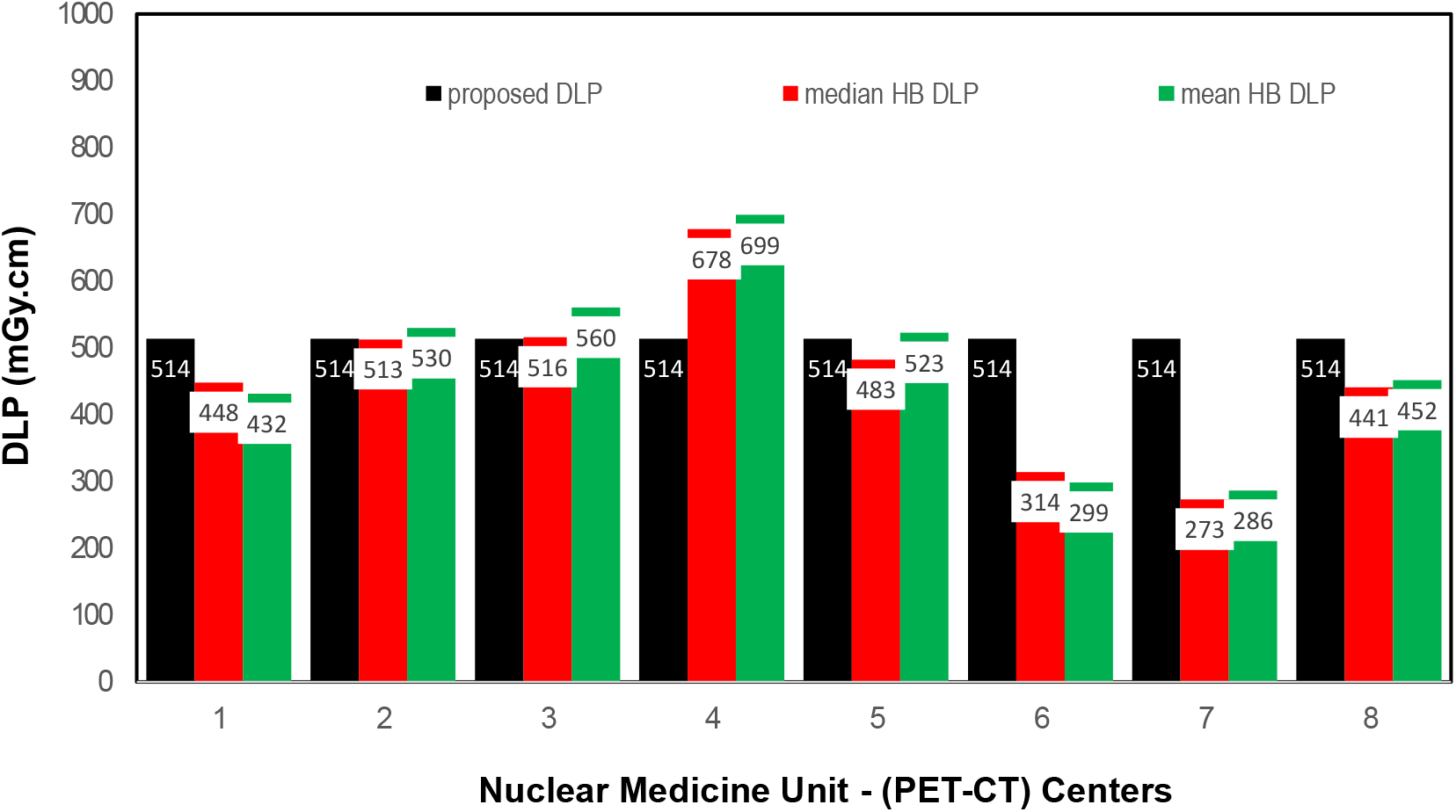
DLP (75^th^ percentile) for each PET-CT unit, compared to the proposed NDLP for HB data.

**Figure 3:**
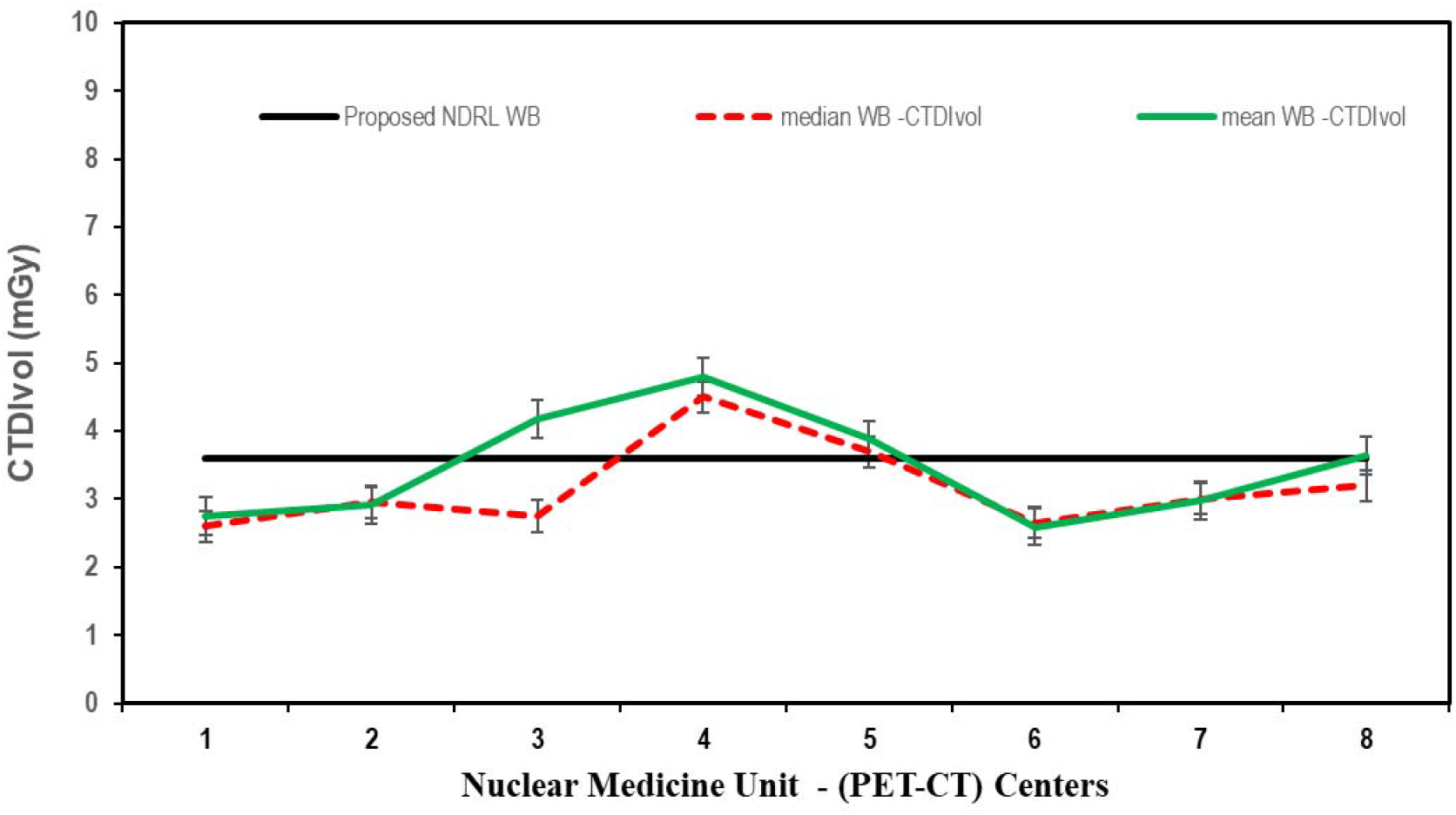
CTDIvol (75^th^ percentile) for each PET-CT unit, compared to the proposed NDRL for WB data.

**Figure 4:**
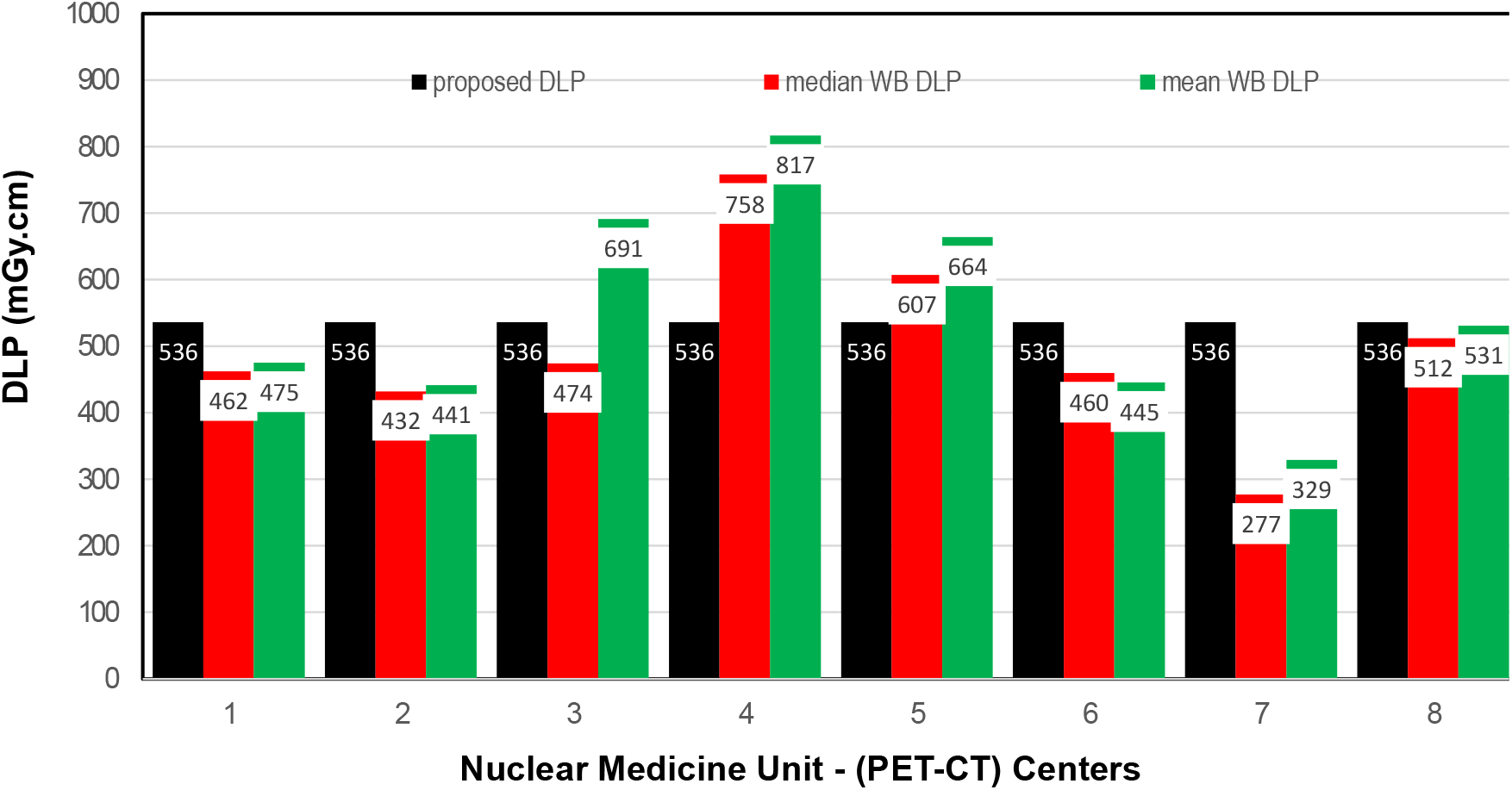
DLP (75^th^ percentile) for each PET-CT unit, compared to the proposed NDRL for WB data.

**Figure 5:**
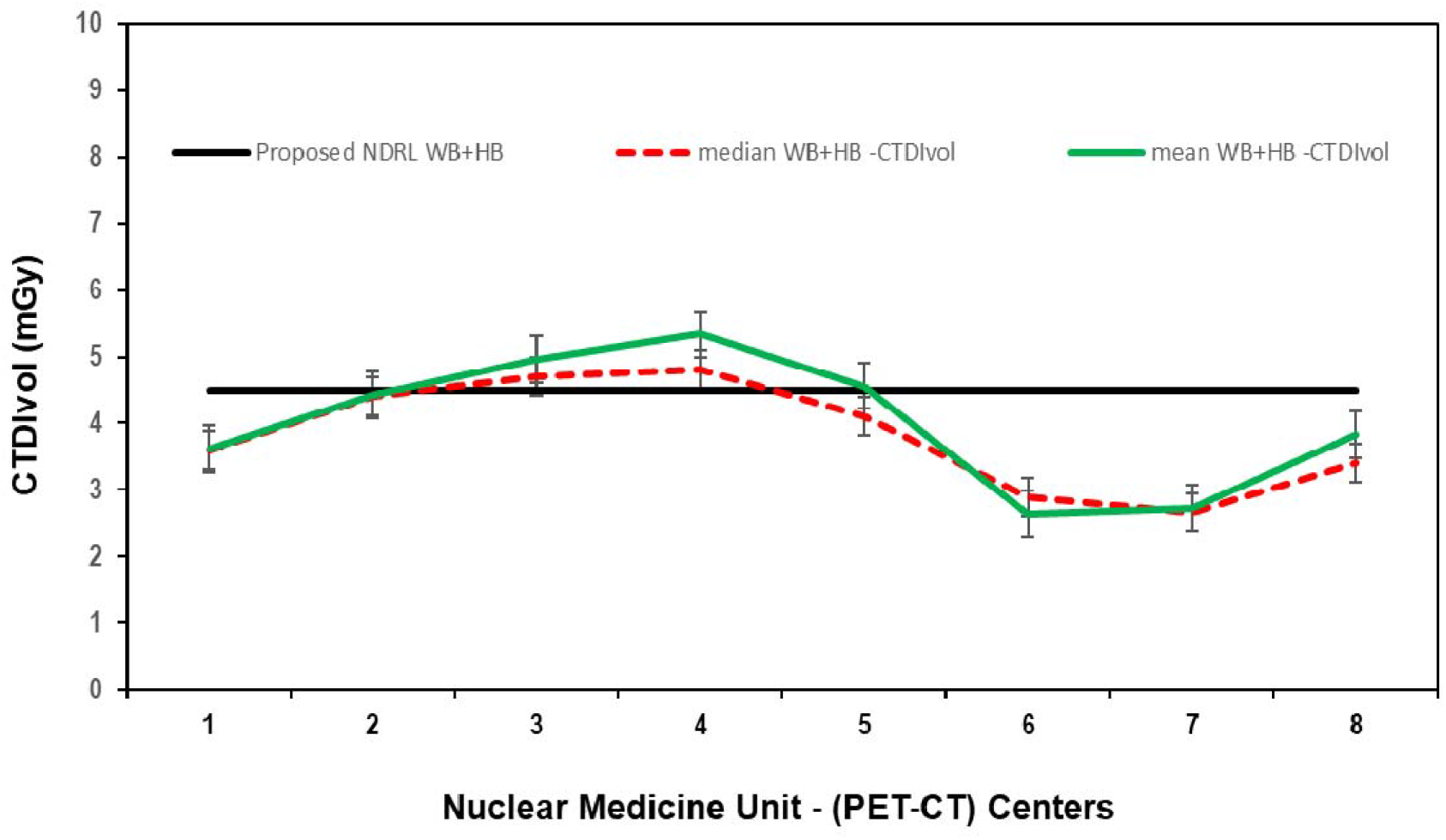
CTDIvol (75^th^ percentile) for each PET-CT unit, compared to the proposed NDRL for WB+HB data.

The ratio of maximum to minimum mean doses for HB and WB scans between different centers for the same clinical studies varied between 2.3-7.3 for HB and 2.1-7.3 for WB. **Figures [7-8]** are presenting trends of CTDIvol and ED variations over 2018 and 2019 with reference to the NDRLs and comparatively. The NDRL for the second phase study (2019) showed an improvement of about 9.1% over the 2018 NDRL result. For 2019, the DRL % deviations from the NDRL varied from [-4% to +40%] for each center.

**Figure 6:**
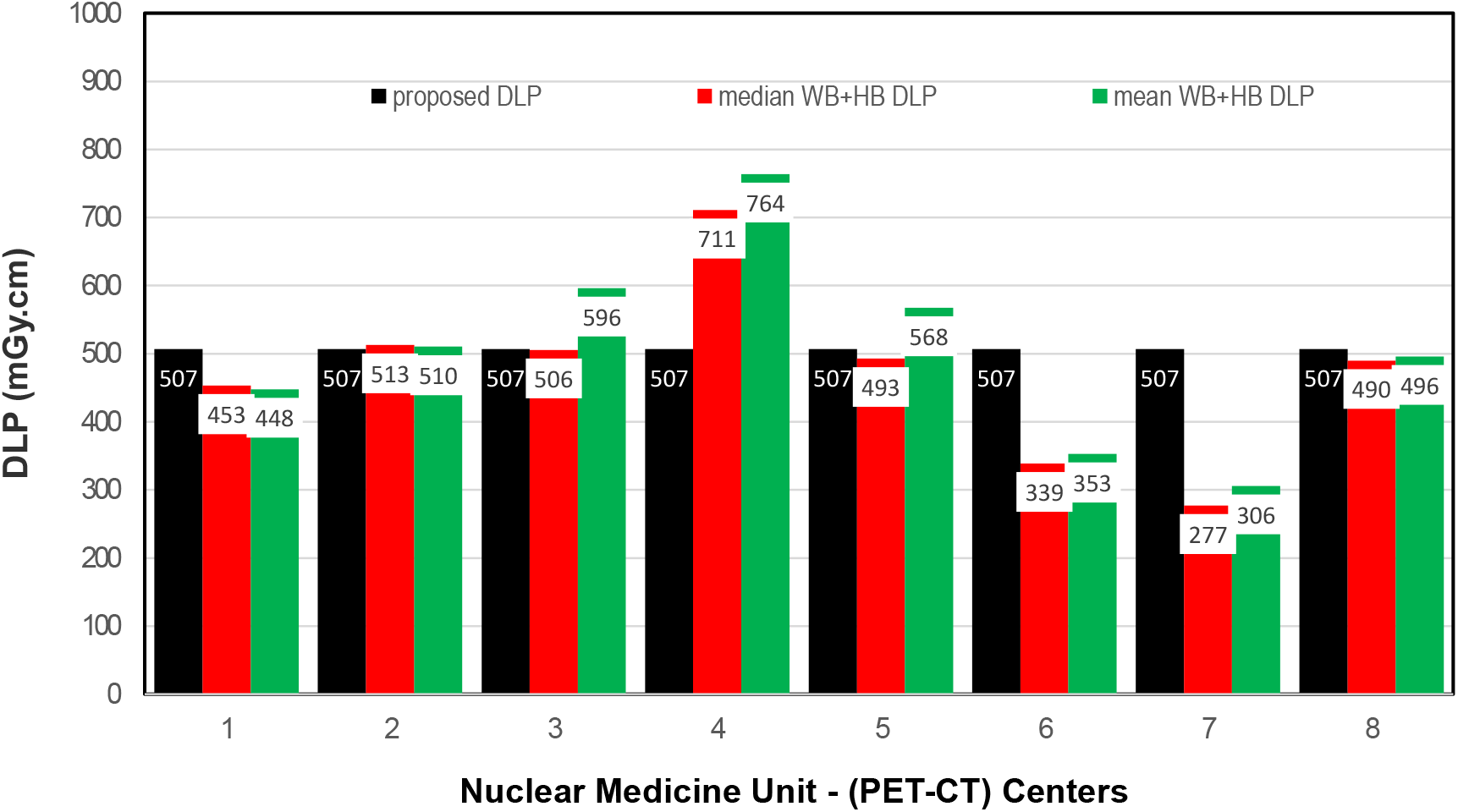
DLP (75^th^ percentile) for each PET-CT unit, compared to the proposed NDRL for WB +HB data.

**Figure 7:**
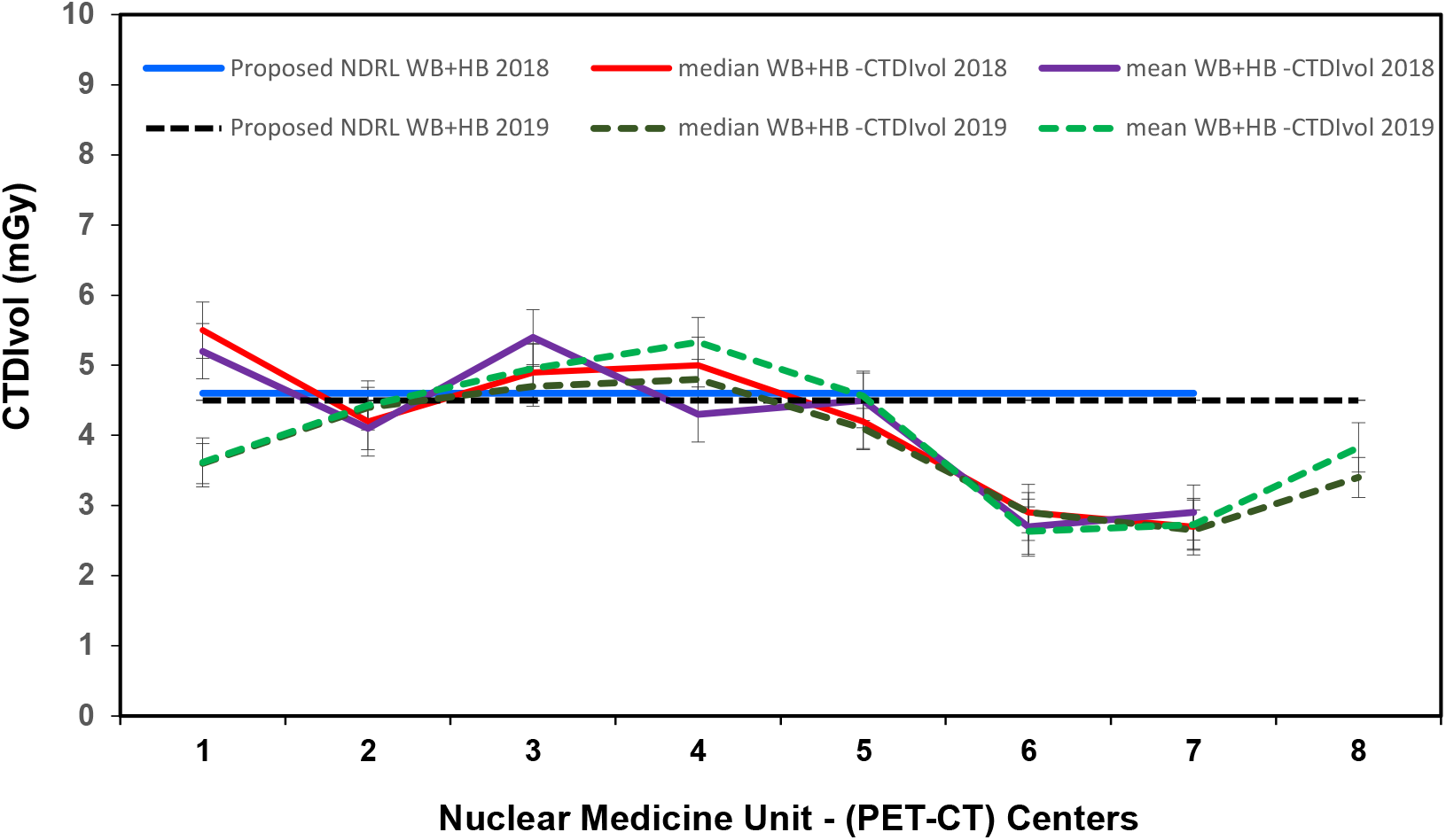
Comparison of CTDIvol (75^th^ percentile) for WB+HB data and proposed NDRL at each PET-CT unit in 2018 and 2019. -ED: Effective Dose

**Figure 8:**
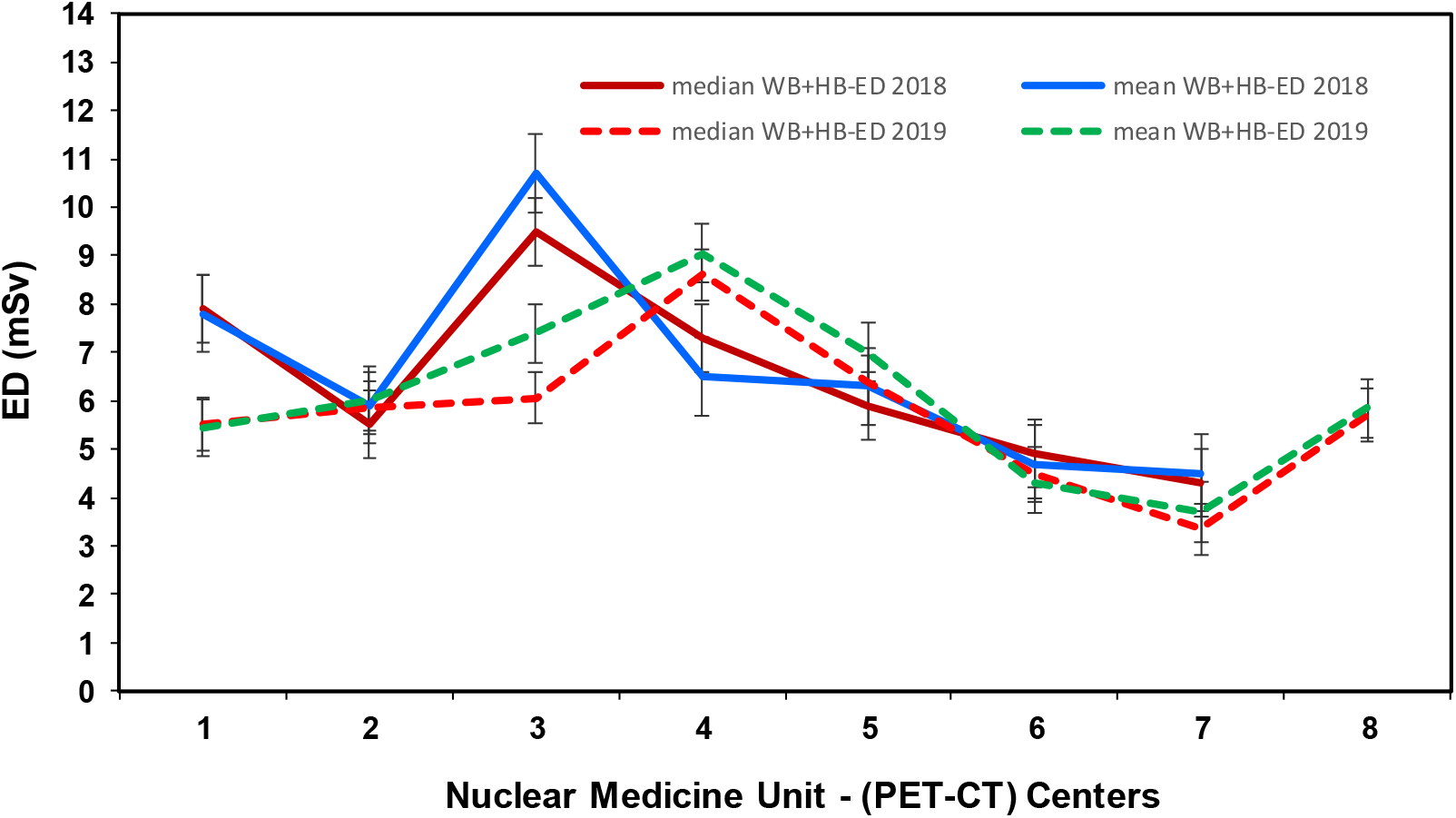
Comparison of CT ED for WB+HB data at each PET-CT unit in 2018 and 2019.

## CONCLUSIONS

It is anticipated that with the establishment of the NDRLs, the Achievable dose and continuous monitoring, it will be possible to optimize practice across the Kuwait and reduce their variations in the next future surveys, and to promote improvements in patient protection and quality care in state of Kuwait. The outcome of this second phase audit has further paved the way to refine the setting of NDRL CT part of the PET / CT examination for Kuwait populations which based on the current facilities and practice that is more realistic than external sites. Equally important, it will facilitate and assists to create a data bank (i.e. National Archive) for the future years to sever as a monitoring tool to elevate quality care for KW populations.

## Data Availability

All presented data in this article are original and are available on request

## DISCLOUSURE

This study was supported by the research grant (PR19-13MN-02) from the Kuwait Foundation of Advancement of Sciences (KFAS) that facilitated the 2^nd^ phase of this investigation which utilized state of art technology, technique and advanced science.

## ACKNOWLEDGEMENTS

The authors would like to thank the following NM PET/CT centers in the state Kuwait for their effective participation in provision of patient CT data for the second consecutive year (arranged alphabetically): Adan Hospital; Al-Jaber Hospital; Chest Diseases Hospital; Cancer Control Center; Farwaniya Hospital; Jahra Hospital; Jaber Al Ahmed Molecular Imaging, and Mubarak Al Kabeer Hospital.

